# Evaluation of Indicators of Reproducibility and Transparency in Published Cardiology Literature

**DOI:** 10.1101/19002121

**Authors:** J. Michael Anderson, Bryan Wright, Daniel Tritz, Jarryd Horn, Ian Parker, Daniel Bergeron, Sharolyn Cook, Matt Vassar

## Abstract

**Background:** The extent of reproducibility in cardiology research remains unclear. Therefore, our main objective was to determine the quality of research published in cardiology journals using eight indicators of reproducibility.

**Methods:** Using a cross-sectional study design, we conducted an advanced search of the National Library of Medicine (NLM) catalog for publications from 2014-2018 in journals pertaining to cardiology. Journals must have been published in the English language and must have been indexed in MEDLINE. Once the initial list of publications from all cardiology journals was obtained, we searched for full-text PDF versions using Open Access, Google Scholar, and PubMed. Studies were analyzed using a pilot-tested Google Form to evaluate the presence of information that was deemed necessary to reproduce the study in its entirety.

**Results:** After exclusions, we included 132 studies containing empirical data. Of these studies, the majority (126/132, 95.5%) did not provide the raw data collected while conducting the study, 0/132 (0%) provided step-by-step analysis scripts, and 117/132 (88.6%) failed to provide sufficient materials needed to reproduce the study.

**Conclusions:** The presentation of studies published in cardiology journals does not appear to facilitate reproducible research. Considerable improvements to the framework of biomedical science, specifically in the field of cardiology, are necessary. Solutions to increase the reproducibility and transparency of published works in cardiology journals is warranted, including addressing inadequate sharing of materials, raw data, and key methodological details.

## Introduction

Cardiology is a field at the forefront of evidence-based medicine. Advances in diagnostic and therapeutic modalities related to imaging, biomarkers, drugs, devices, and minimally invasive procedures have improved clinical outcomes and quality of life.^1^ However, disagreements exist between the results from randomized clinical trials and observational studies – two study designs that often provide the evidentiary base for clinical practice guidelines in cardiology. In such cases, some authors have argued that “evidentiary policies should emphasize the open access to clinical data [and] the responsible analysis of these data by agencies immune to the influence of partisan stakeholders”^2^ to better account for disparate study findings and to improve reproducibility. Take as an example the United States Preventive Services Task Force’s (USPSTF) and American Heart Association’s (AHA) clinical practice recommendations for aspirin prophylaxis to prevent cardiovascular disease. The original recommendations were based on a meta-analysis of five studies that found a 28% decrease in coronary disease, with no improvement in total mortality, and an increased risk of bleeding.^3,4^ A more recent meta-analysis of 11 studies of aspirin for primary prevention found no reduction in mortality and a 0.6% increase in major bleeding.^5^ This most recent analysis lead to a reversal in the clinical recommendations for patients >70 years old and called into question its use for patients between 40 and 70 years of age.^6^ One possible mechanism to prevent similar reversals of practice in cardiovascular medicine is to advocate for reproducible, transparent research practices.

Concern over the lack of reproducibility in scientific research has been well documented.^7–10^ Attention surrounding this issue has grown to such prominence that a recent survey in Nature reported that 90% of respondents from the scientific community agreed that a ‘reproducibility crisis’ exists.^11^ The National Institutes of Health (NIH) recently called for “immediate and substantive action” to address this shortcoming.^12^ This call to action stems from reports that only 10–25% of scientific experiments are reproducible,^8,11,13^ resulting in $28 billion per year spent on irreproducible preclinical research in the United States alone.^14^ In 2018, the National Academies of Science, Engineering, and Medicine drafted a congressionally-mandated report, Reproducibility and Replicability in Science^15^, which outlines recommendations for research stakeholders – including researchers, journals, funding agencies, and academic institutions – to improve the reproducibility and transparency of scientific research. Prior to release of the National Academies’ report, this body, referred to as the Institute of Medicine at the time, established the Committee on Strategies for Responsible Sharing of Clinical Trial Data that recommended all studies with a novel study design provide detailed data sharing, metadata, protocols, statistical analysis plans, and analytic code to be considered reproducible.^16,17^

Within the cardiology specialty, some journals have added requirements for authors to share data and append supplementary materials to published reports in an attempt to improve reproducibility. For instance, the American Heart Association journals all adhere to Transparency and Openness Promotion (TOP) Guidelines that outline mechanisms for sharing data, statistical code, and other materials to facilitate reproducibility.^18^ The urgency for reproducible research highlights the need for closer examination of practices within the field of cardiology. In this study, we evaluate eight indicators of reproducibility and transparency posited by Hardwicke et al.^19^ in a random sample of studies published in cardiology journals. Results from this investigation showcase areas where improvements are most needed and serve as baseline data for future, comparative investigations.

## Methods

This investigation employed an observational, cross-sectional design. Since the Open Science Framework has generated a new infrastructure that advocates for research transparency, we have supplied our complete protocol, raw data, and other necessary materials at https://osf.io/x24n3/. Using a similar methodology and protocol to that outlined by Hardwicke et. al.^19^, with slight modifications, our study focused, specifically, on the field of cardiology. This study was not subject to institutional review board oversight according to the U.S. Department of Health and Human Services’ Code of Federal Regulation 45 CFR 46.102(d) and (f)^20^ because our study lacked human subject participation. The present investigation was reported using the guidance for conducting meta-research outlined by Murad and Wang.^21^ In addition, the Preferred Reporting Items for Systematic Reviews and Meta-Analyses (PRISMA) guidelines were used when appropriate. Our primary endpoint was to assess for the presence of eight indicators that support transparent research practices, allowing for feasible reproduction of cardiology studies.

### Journal and Study Selection

For this project, one of us (DT) searched all journals in the National Library of Medicine (NLM) catalog using the subject term “Cardiology[ST]”. Our search was conducted on May 29, 2019. To meet inclusion criteria, the journals must be in English and MEDLINE indexed. The list of journals generated from our search of the NLM catalog was then extracted by electronic ISSN number (or the linking ISSN if the electronic version was not available). The journal search string of ISSN numbers was then used in PubMed (which encompasses the entire MEDLINE database) on May 29th, 2019 to gather all publications. For our final list of publications, we included only those from January 1, 2014 to December 31, 2018. Using Excel’s random number function, a random number was assigned to all studies. A random sample of 300 studies were chosen for our analysis. This list of studies is available on the Open Science Framework (https://osf.io/4mtwq/). To ensure a wide range of publications, we did not restrict any particular study designs from inclusion.

### Extraction Training

Prior to data extraction, two investigators (MA, BW) completed thorough training to establish consistent study evaluation methods between investigators. Training consisted of an in-person session that reviewed study objectives, design, protocol, the Google form used for data extraction, and how to obtain the pertinent information from sampled publications. The methodology was demonstrated by DT using two example studies. MA and BW then independently extracted data from three sample studies in a blinded, duplicate manner. Following this mock data extraction, the pair reconciled all differences between them. Applying the same blinded, duplicate data extraction process, MA and BW extracted data from the first 10 publications of the list generated using the NLM catalog. This was followed by a final consensus meeting to amend all discrepancies. This initial training session was recorded and available online for reference (https://osf.io/tf7nw/).

### Data Extraction

Upon completing training, these investigators extracted data from the remaining 290 publications in a blinded, duplicate fashion. Data extraction began on June 3, 2019 and was completed on June 10, 2019. Once data extraction was complete, a final consensus meeting was held by MA and BW to resolve any discrepancies. If necessary, DT was available for adjudication. We divided publications into two separate categories: (1) publications consisting of empirical data (e.g. clinical trial, cohort, case series, case reports, case-control, secondary analysis, chart review, commentary [with data analysis], and cross-sectional) and (2) studies lacking empirical data (e.g., editorials, commentaries [without reanalysis], simulations, news, reviews, and poems). Using a pilot-tested Google form similar to that created by Hardwicke et al.^19^, with additions, investigators were prompted to identify whether a study had important indicators considered necessary to reproduce a study (https://osf.io/3nfa5/). The Google form used to extract data prompted investigators to extract appropriate information based on the individual study design of each publication. Studies lacking empirical data were only analyzed for the presence of funding source(s), disclosure of conflicts of interest, open access, and impact factor (2017, 2018 and five-year impact factor). Case reports and case series were extracted using the same methods as studies that lacked empirical data. Case reports and case series typically lack pre-specified protocols and are, therefore, inherently difficult to reproduce.^22^ For systematic reviews/meta-analyses, all indicators were evaluated with the exception of materials availability since it is not expected for them to include additional materials. Our form also expanded the extraction constructed by Hardwick et al. to include the following options for study design: case series, cohort, secondary analysis, systematic review/meta-analysis, chart review, and cross-sectional studies. Finally, the options for funding source were expanded to include more specific identifiers of funding sources that included: university, hospital, public, private/industry, and/or non-profit.

### Assessment of Open Access Data

To assess the public’s ability to access each publication included in our sample, we attempted to obtain the full-text PDF version of each publication via a systematic process. First, we searched for publication title, DOI, and/or PubMed ID using Open Access Button (https://openaccessbutton.org/). If this search method was unsuccessful, investigators then searched Google Scholar and PubMed for publication title, DOI, and PubMed ID. If the full-text was still unobtainable after all search methods were exhausted, the publication was considered to be paywall restricted and, therefore, not accessible to the public.

### Evaluation of Replication and Whether Publications Were Included in Research Synthesis

For studies with empirical data, we searched Web of Science to determine whether the present study: (1) was included as part of a replication study in subsequent publications and (2) had been cited in systematic reviews and/or meta-analyses. By reviewing the titles and abstracts of all publications in which the study was cited, we made the determination of the frequency with which the given study was cited in replication studies or systematic reviews/meta-analyses. This process was performed using a blinded, duplicate data extraction process.

### Analysis Plan

Outcomes are presented as percentages with 95% confidence intervals. We reported descriptive statistics for each category using Microsoft Excel.

## Results

Our NLM search identified 280 journals for cardiology. Only 150 journals met the inclusion criteria and yielded 145 471 publications from 2014 to 2018. A random sample of 300 publications yielded 290 eligible publications and 10 inaccessible ones (Figure 1). The variables analyzed depended on individual study designs. Thus, Supplementary Table 1 outlines a detailed description of the included publications and the corresponding analysis of the eight queried indicators of reproducibility.

**Figure 1:**
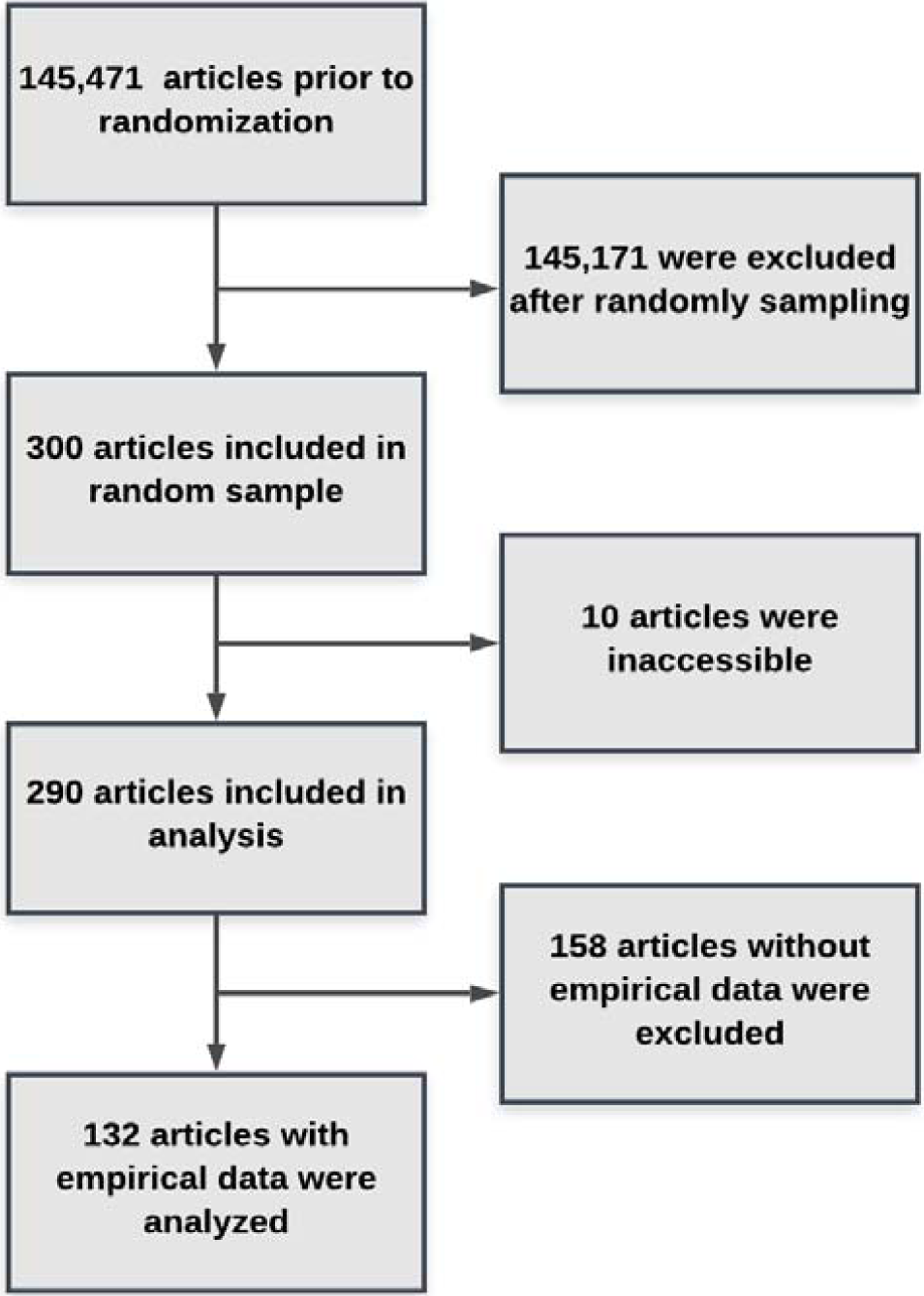
Flow Diagram for Included and Excluded Studies.

### Sample Characteristics

Our final analysis included publications lacking empirical data (n = 128), including editorials, commentaries (without reanalysis), simulations, news, and reviews. Even though case reports and case series (n = 30) provide empirical data, the given methodologies are typically either insufficient for reproduction or absent (as detailed by Wallach et al.^22^). We, therefore, extracted these study designs in an identical fashion as studies with non-empirical data. The common absence of reproducible methodology in case reports and case series formulated the basis for excluding these study types from certain analyses. Thus, our final sample size consisted of 132 publications with empirical data from reproducible study types (e.g. cost-effectiveness/decision-making, clinical trial, cohort, case-control, secondary analysis, chart review, commentary [with data analysis], survey, laboratory, and cross-sectional). Studies lacking empirical data made up the largest percentage (44.1%, 128/290) of studies included in our sample. The median five-year journal impact factor was 4.023 (range: 0.05-23.425). Journal impact factors were inaccessible for 16 publications (Supplemental Table 2). Additional reproducibility and transparency characteristics for our sample of publications from cardiology journals are available in Table 1 and Supplemental Table 3.

**Table 1.**
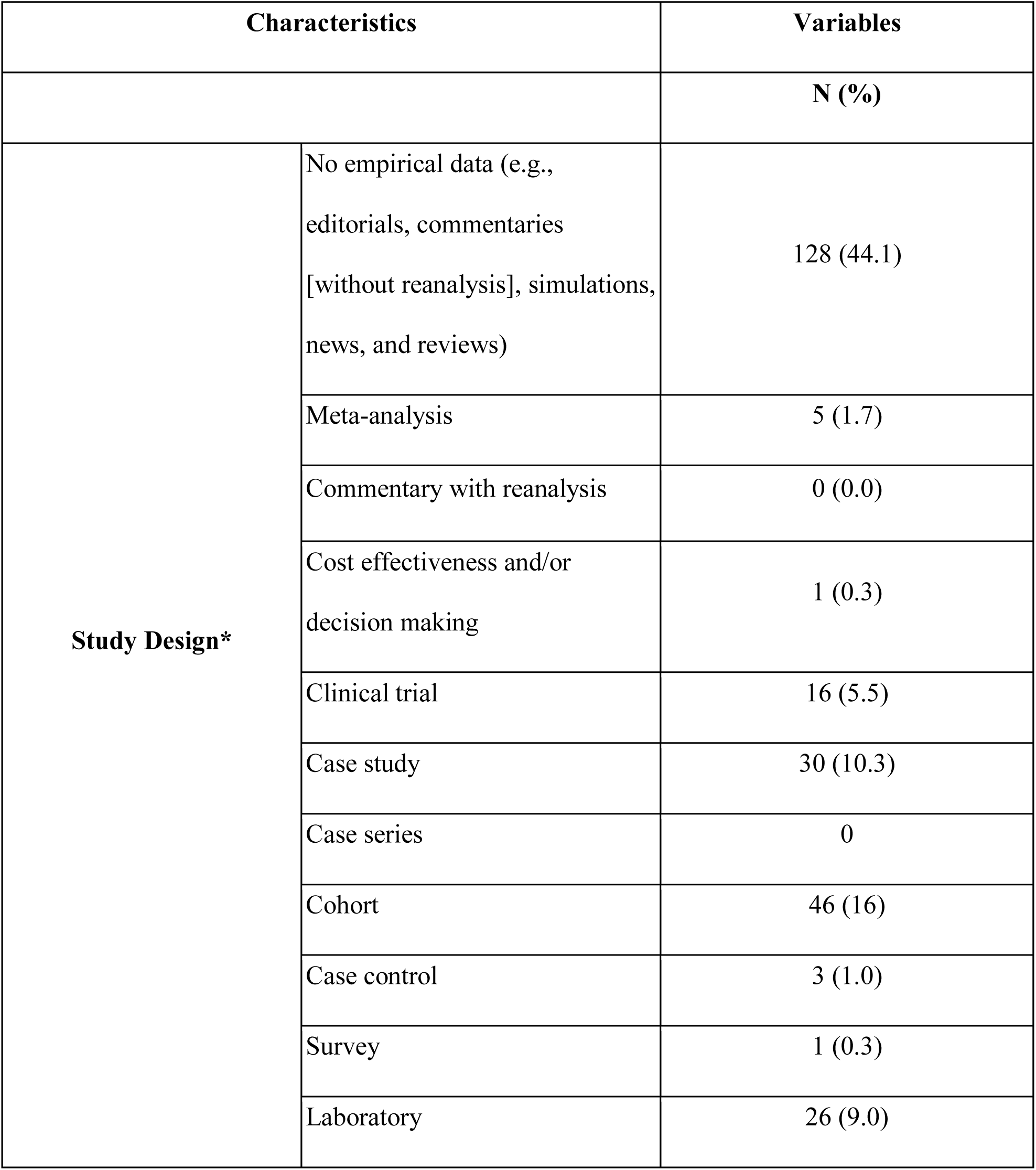
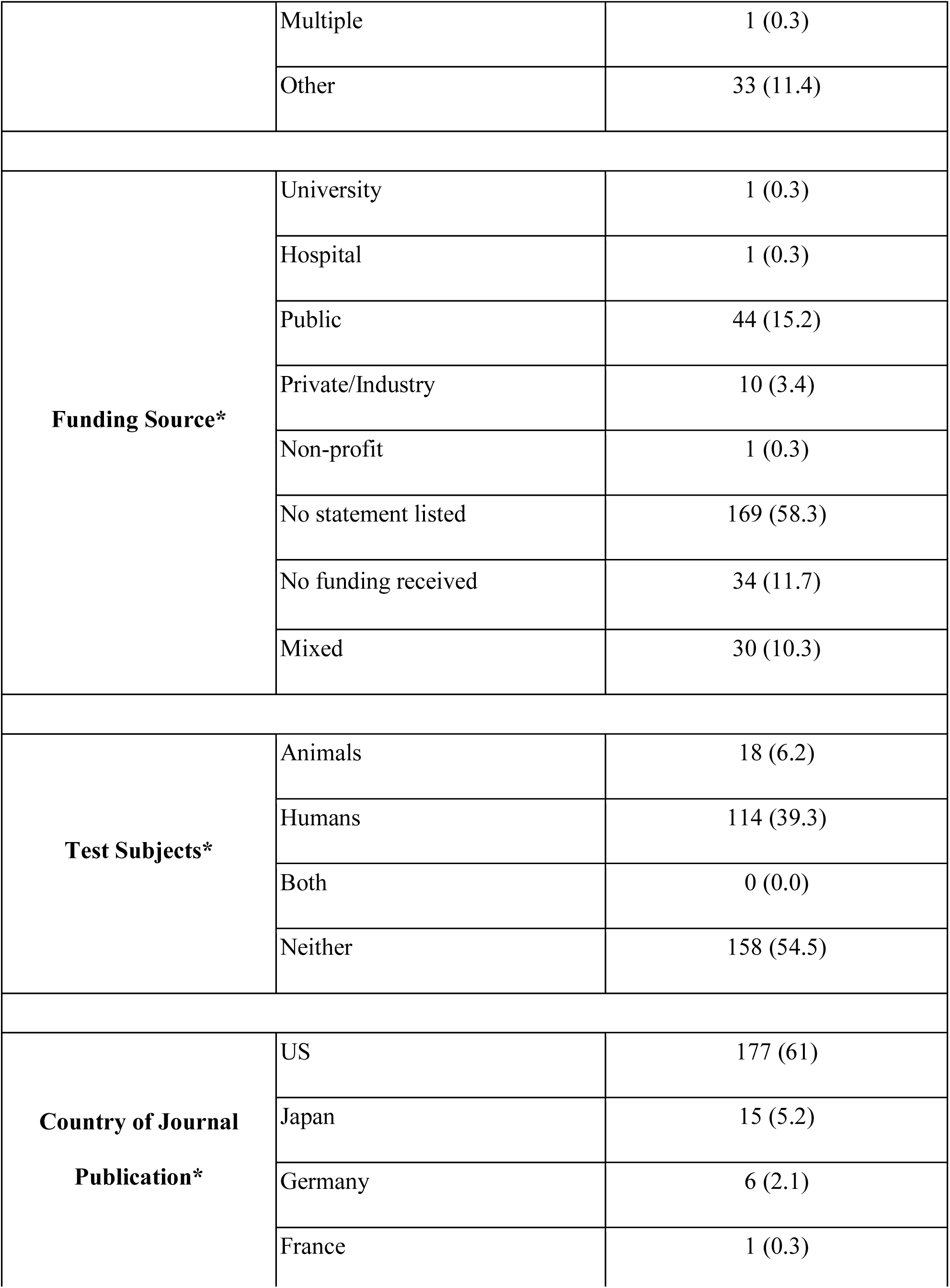
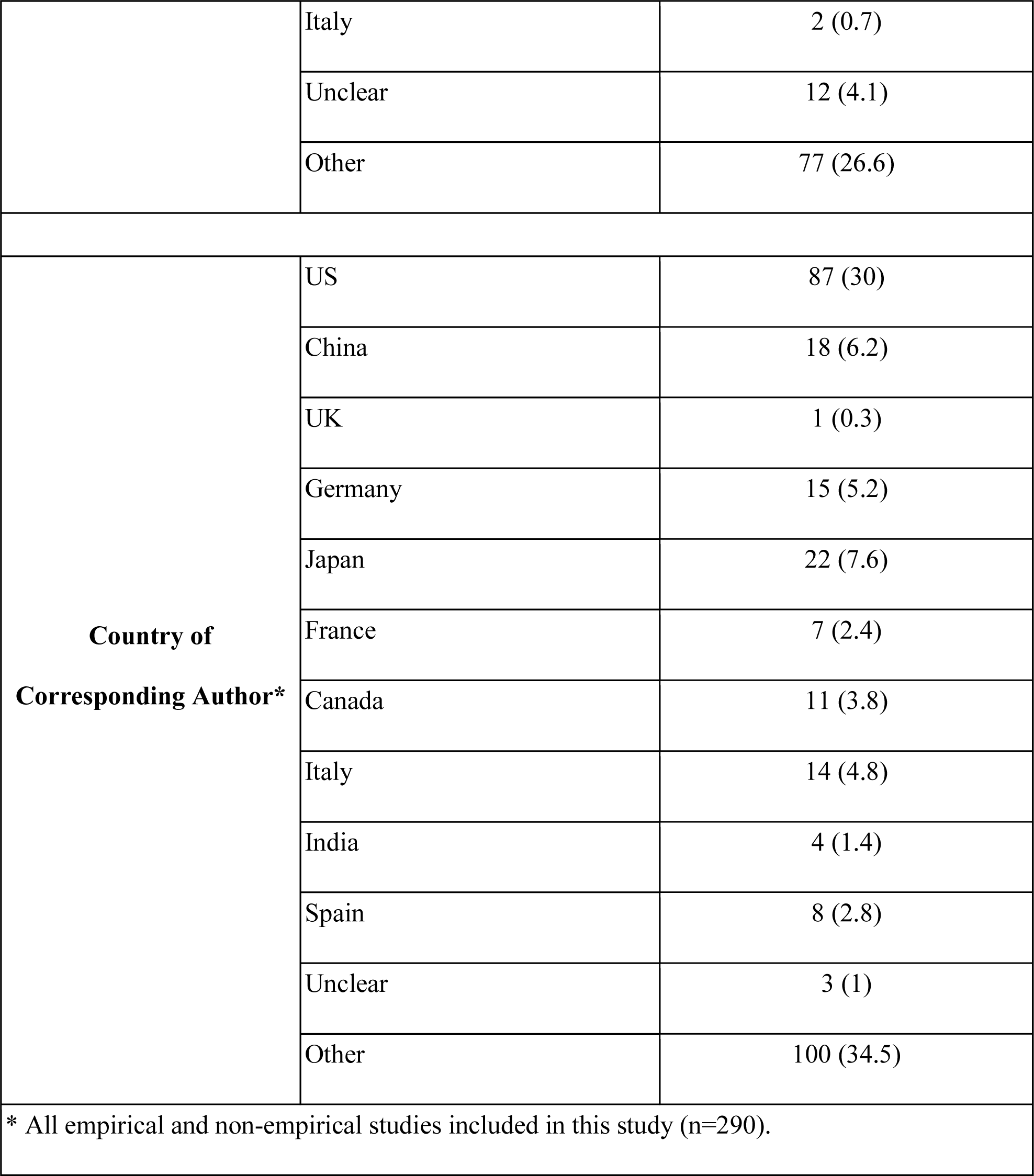
Reproducibility and Transparency Characteristics for Sample of Publications in Cardiology Journals.

### Article Availability

Among the 290 accessible publications, 168 (58%) were publicly available, whereas 122 (42%) were only available through a paywall (Supplemental Table 3). Furthermore, we considered the 10 publications for which full-text was unavailable to be paywall restricted. Thus, 132 publications (of 300, 44%) were considered to be restricted behind a paywall.

### Materials Availability

We considered “materials” and “protocol” as any study item and/or document that might be deemed necessary to accurately reproduce the study. Our analysis revealed only 15 publications (of 132) containing empirical data that provided a materials availability statement, and only one publication (of 132) that explicitly stated no supplemental materials were available (Supplemental Table 3). Author statements declaring how to obtain supplemental materials included online journal supplemental appendices (n = 10), an online third party (n = 2), requesting from the corresponding author (n = 2), and a personal or institute website (n = 1). Supplemental materials that authors claimed to be available were only accessible for only seven (47%) of the 15 publications. We did not attempt to verify the availability of materials by requesting them from the corresponding author.

### Protocol Availability

In addition, only four publications (of 127) with empirical data – excluding case reports, case series, and meta-analyses, as detailed in the methods section – provided links to accessible protocols. Of the four publications claiming to provide access to full protocols, two of the links did not successfully navigate to an accessible protocol. The remaining two publications linked to accessible methods and analysis plans (Supplemental Table 3).

### Data Availability

Complete and accurate validation of analyses performed within scientific literature requires access to the raw data collected by a particular study. For the purpose of our study, “raw data” refers to a detailed record of all collected information in an unmodified, digital form (e.g., sampling units of testing participants). Ninety-nine publications containing empirical data lacked data availability statements with the remaining 33 publications containing a data availability statement. Of the 33 publications with data availability statements, 28 of the referenced supplementary materials were available via the associated online journal, three upon request from the corresponding author(s), and one via an online third party repository (Supplementary Table 3). During data extraction, we were only able to access, download, and open supplementary data files for 19 of the 33 publications claiming additional data availability. Furthermore, only six of the 19 publications with accessible supplementary data provided all raw data necessary to reproduce all calculations and statistical analyses performed by the researchers in the given study. From our sample, less than 4% (6/132) of publications in cardiology journals provide access to complete records of raw numerical data. We did not attempt to verify the 3 publications which claimed to provide data upon request from the corresponding author.

### Analysis Script Availability

Our definition of analysis script encompassed a detailed description of data preparation and either step-by-step instructions for using point--and--click software, analysis code (e.g., Python, Mathlab), or syntax (e.g., SPSS, Stata). Our analysis failed to identify a single publication (0 of 132) within our sample that included an analysis script statement.

### Preregistration

Open access registries, such as ClinicalTrials.gov (a public database created by the Food and Drug Administration [FDA] and the National Institutes of Health [NIH]), provide specific information pertaining to a given study, including hypotheses, methods, and/or analysis plans.^23^ Less than 5% of publications (6 of 132) with empirical data provided information regarding registration prior to the commencement of the study. In addition, only four of the six provided sufficient information to completely access the study registration. Three preregistered studies (of four, 75%) provided study methods, two (of four, 50%) provided hypotheses, and two (of four, 50%) provided analysis plans (Supplemental Table 1).

### Conflict of Interest and Funding Statements

Among the 290 publications, 241 (83%) provided a conflict of interest (COI) statement. Sixty-four publications (22%) stated the authors had no COI, whereas 177 publications (61%) disclosed authors having one or more COI. Forty-nine publications (17%) did not include a conflict of interest statement. A total of 168 publications (58%) did not include a funding statement, and 34 publications (12%) reported the study was conducted without funding. Of the 122 publications reporting a funding source, the majority (n = 71) of the funds were provided by public sources (Table 1).

### Replication and Evidence Synthesis

Findings from previous studies may be included in future systematic reviews and meta-analyses that can be used to help answer focused clinical questions^24^; however, our analysis shows the publications included in our sample were not commonly cited in other studies. A total of 26 publications (out of 132, 19.7%) with empirical data were cited in one or more subsequent meta-analysis or systematic review (range: 1 to 7; Supplemental Table 4). Our sample did not include a single self-identified replication study.

## Discussion

The climate of studies published in cardiology journals does not appear to facilitate reproducible research. We found that only 25% of publications included data availability statements and only 11% included material availability statements. Despite some publications providing additional data and materials, less than 5% provided the complete raw data used to calculate study findings. Furthermore, we found that publications in cardiology journals do not consistently provide analysis scripts and step-by-step protocols to thoroughly reproduce a study.

Variable analytical approaches to the same data set makes it difficult to reach the same or similar outcomes.^25^ Incomplete analysis scripts allow replication studies to employ different analytical approaches with the same data set, which may result in different outcomes.^26^ This lack of transparency makes it difficult to verify the accuracy and legitimacy of results. Our findings are similar to a previous study examining reproducibility in biomedical research that concluded certain indicators of reproducibility – including data sharing, materials availability statements, and disclosures of funding sources – are inconsistently reported.^22^ The reproducibility surrounding biomedical research will continue to be suboptimal if open access to published literature remains restricted, and the necessary components of a study are not provided, such as all supplemental materials, raw data, and key methodological information.

The majority of studies included in our analysis failed to provide access to all raw numerical data, suggesting a lack of data sharing in cardiology studies, a well-documented obstacle to reproducible research. For example, a survey in Nature addressing inaccessible raw data from the original laboratory as a barrier to reproducibility reported that nearly 70% of scientists were unable to successfully reproduce study outcomes in the medical literature.^11^ The idea of inferential reproducibility (e.g. rerunning the analyses using the same raw data from the original study), focuses on an essential aspect of research reproducibility. This idea is based upon the notion that an individual should be able to start with the same raw data and end with similar conclusions; however, it has been previously demonstrated that different conclusions may be reached following analysis of the same raw data sets.^27,28^ For example, a 2018 study concluded a “significant variation in the results of analyses of complex data may be difficult to avoid, even by experts with honest intentions.” ^25^ The practice of data sharing has the potential to increase reproducibility and transparency in biomedical research; however, providing sufficient protocols and analysis scripts used in data analysis is equally important.

Failure to share sufficient protocols and analysis scripts to rerun computations contributes to the non-reproducibility and lack of transparency in research. For example, a 2018 study^28^ demonstrated that researchers often repeat a previous experiment using the same methodological steps and protocols to determine the level of reproducibility.. Inadequate methodological reporting and absent analysis scripts makes it difficult to reproduce study outcomes and highlights claims made in previous studies regarding the poor reproducibility of research.^9,10,13^ However, it should be noted that another barrier to reproducing scientific literature is a result of digital information becoming “unusable, inoperative, or unavailable because of technological breakdown and evolution or lack of continued curation.” ^29^ Given that the information necessary to reproduce a study is frequently housed within the published text, the lack of open access to medical literature can also perpetuate non-reproducibility.

Studies available via open access have a greater chance of being subjected to criticism from a broader audience; therefore, lack of open access is a barrier to reproducibility. The inability to view original research materials and protocols in their entirety precludes a comprehensive evaluation of a study. ^30^ For example, the authors of a 2017 study credit the lack of open access as contributing to the inability to obtain full protocols and analysis scripts.^31^ In a sense, open access provides another means of transparency for those who would seek to reproduce a study and validate outcomes. Nearly one-half of the publications included in our sample were inaccessible due to paywall restrictions. This is comparable to a previous study investigating reproducibility in biomedical sciences which found that approximately 43% of publications with a PubMed ID were not publicly available.^22^ Preregistration of studies might help overcome this by making accessible hypotheses, protocols, primary outcome measures. Preregistering studies on public registries may provide sufficient information to reproduce published and unpublished research alike, even if the full-text is not publicly available.^32^

### Future Directions

The optimum framework by which to address the obstacles surrounding non-reproducibility of studies in cardiology journals is unclear. Requiring authors at the time of publication to submit all raw data and materials necessary to conduct the study might increase the ease with which computations can be reproduced and validated. Efforts have been made to increase the rate at which data is openly shared by authors. For example, the TOP initiative, a 2015 promotion pioneered by Nosek and colleagues ^32^, proposes guidelines on data sharing to help increase the dissemination of research data while simultaneously helping increase the reproducibility of scientific literature. Many journals, including Science, implement aspects of the TOP guidelines to hold authors to a higher standard by requiring disclosure of all data used in the analysis such that any researcher may reproduce the findings.^31^ Additional journals implementing these same TOP guidelines include the American Heart Association and American Stroke Association journals.^33^ More widespread TOP guideline adoption has the potential to minimize the reproducibility crisis. Furthermore, reviewers have taken the initiative to increase the clarity of scientific literature by pledging to adhere to The Peer Reviews’ Openness Initiative. By making this pledge, reviewers agree to refuse a comprehensive review of any manuscript from authors who fail to make raw data openly accessible without sufficient reason to the contrary.^34^ For these reasons, we urge all journals to consider adopting similar data sharing mandates to make raw data available from all publications, helping to address one cause of non-reproducibility.

While we support initiatives to increase the amount of openly accessible information through data and materials sharing, we also acknowledge the potentially problematic issues that may occur if more raw data is openly available. For example, Lango and Drazen^35^ warn that data sharing has the potential for readers to misinterpret or not completely understand the means by which data were collected. Furthermore, they also assert that data sharing allows for the opportunity to drastically increase the number of “Research Parasites” – researchers who had no part in the original collection, analysis, and interpretation of data, but instead use another person’s data for self-gain. Adopting a symbiotic method of data sharing might be a compromise for the research community’s needs. A possible solution has been proposed by Lango and Drazen^35^ to permit collaborative data sharing between researchers. This proposal suggests that when a novel idea is posited by a second party researcher, the novel study designer should then locate potential collaborators that have already completed the groundwork data collection. Using the collaborator’s data, both parties work alongside one another on the novel experiment. Not only would this solution ensure accurate representation of previously collected data, but conducting research in this manner may facilitate appropriate co-authorship for all who contributed to the novel investigation. Regardless of the methodology by which data is shared, more transparent and openly accessible information to reproduce any given study is necessary.^35^

In addition, it has been argued that all studies should be preregistered prior to commencement of the study in hopes of increasing transparency in medical research. For example, Munafo et al.^31^ suggests the creation of a system whereby the study is preregistered and makes publicly available the following: results, study design, primary outcome, and analysis plan. We agree with the aforementioned proposal, since preregistration might help mitigate analytical manipulation, such as outcome switching and p-hacking – misrepresenting data and/or statistical analyses in such a way that a non-significant finding is found to be significant.^36^ By requiring a more robust standard of reporting analysis plans and step-by-step protocols through preregistration, future researchers might avoid difficulties when trying to replicate a study.

Taking steps to address this concern, The Center for Open Science provides badges to publications which are preregistered, include raw data, and all necessary materials.^31^ Badges signify the importance these journals place on information availability; however, the influence these badges have on open science practices is not well understood and has been the subject of debate.^37^ Despite the unknown influence these badges have on the dissemination of medical research, we encourage a more widespread application of incentive-based guidelines with the goal of increasing the availability of study protocols, materials, and data, even for publications not openly accessible.

### Strengths and Limitations

Our study has many strengths but is not devoid of limitations. The use of open science practices (making available all materials, protocols, analysis plans, and raw data) allows for greater transparency and reproducibility of our study. The blinded, double extraction technique implemented – the gold standard for meta-research data extraction^38^– increases the reliability of our findings. In addition, the robust training provided to investigators prior to initiating this study helped to ensure the reliability of the study results. Despite employing the gold standard to ensure the reliability of data extraction, one limitation of our study includes the possibility that supplemental materials, data, protocols, analysis scripts, and other additional information went unobserved, thereby affecting the outcomes of our study. Second, our analysis of each publication was limited to what was available to investigators through PDF versions and any listed supplemental materials. Additional information necessary to reproduce a study might be obtainable through correspondence with the primary author; however, we did not attempt to obtain information via such methods. Lastly, the results from this cross-sectional analysis of cardiology publications may not be generalizable to other medical specialties. For these reasons, careful interpretation of our findings as a lower bound estimate of study reproducibility in cardiology journals is warranted.

In conclusion, considerable needs exist regarding the reproducibility of studies in cardiology. Current data sharing, disclosure of materials and analysis scripts, as well as access to step-by-step protocols remain unacceptably low. By failing to make essential aspects of scientific literature available, the reproducibility crisis surrounding biomedical research will continue. To promote further transparency and reproducibility, recommendations are proposed to the editors and to the framework of requirements for article submission in cardiology journals to surmount these shortcomings.

## Data Availability

Since the Open Science Framework has generated a new infrastructure that advocates for research transparency, we have supplied our complete protocol, raw data, and other necessary materials at https://osf.io/x24n3/.

## Affiliations

From Oklahoma State University Center for Health Sciences, Tulsa, Oklahoma, USA (J.M.A, B.W., D.T., J.H., M.V.); From Oklahoma State University Medical Center - Department of Cardiology, Tulsa, Oklahoma, USA (I.P., D.B., S.C.)

## SUPPLEMENTAL MATERIALS

**Supplemental Table 1.**
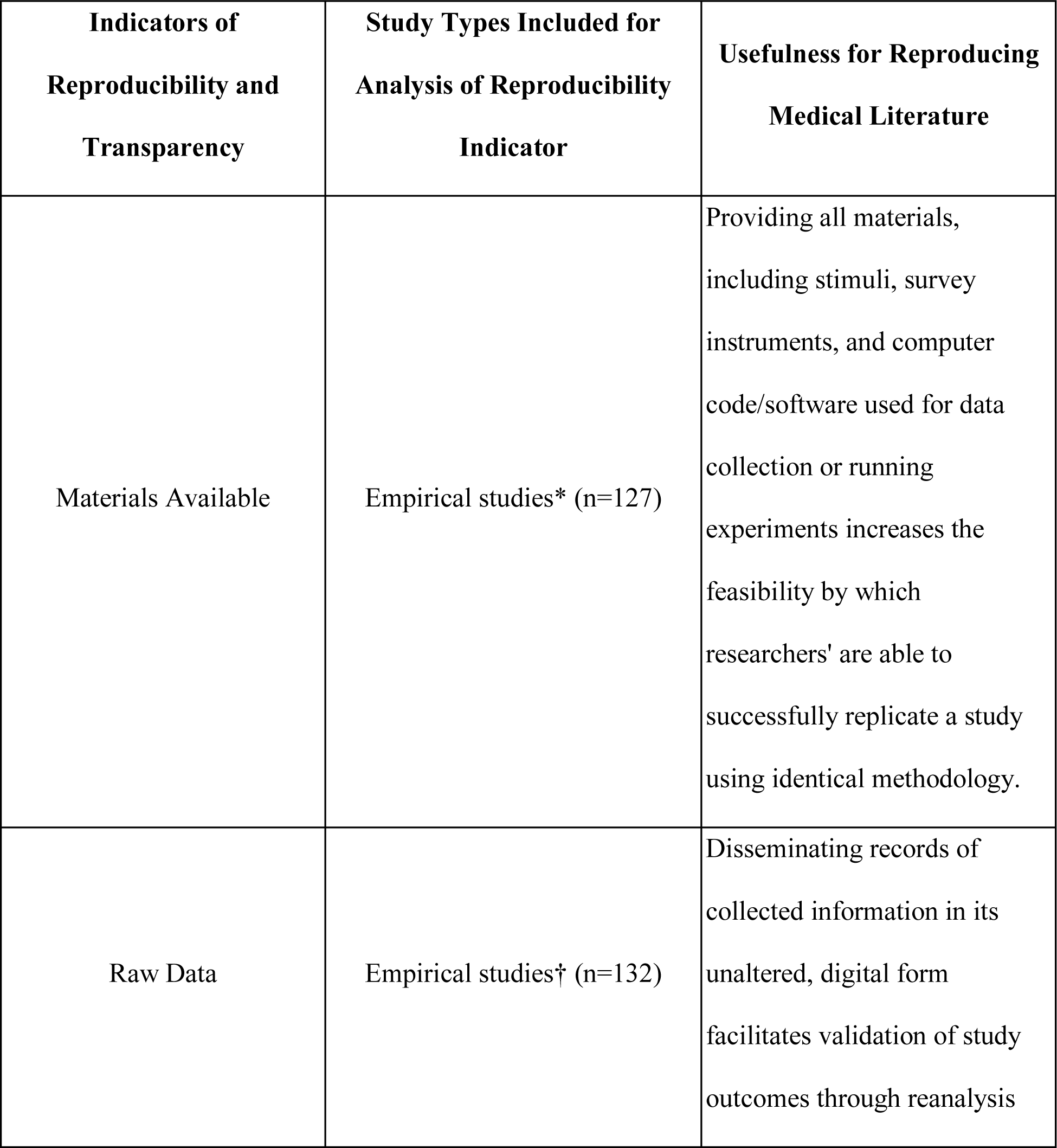
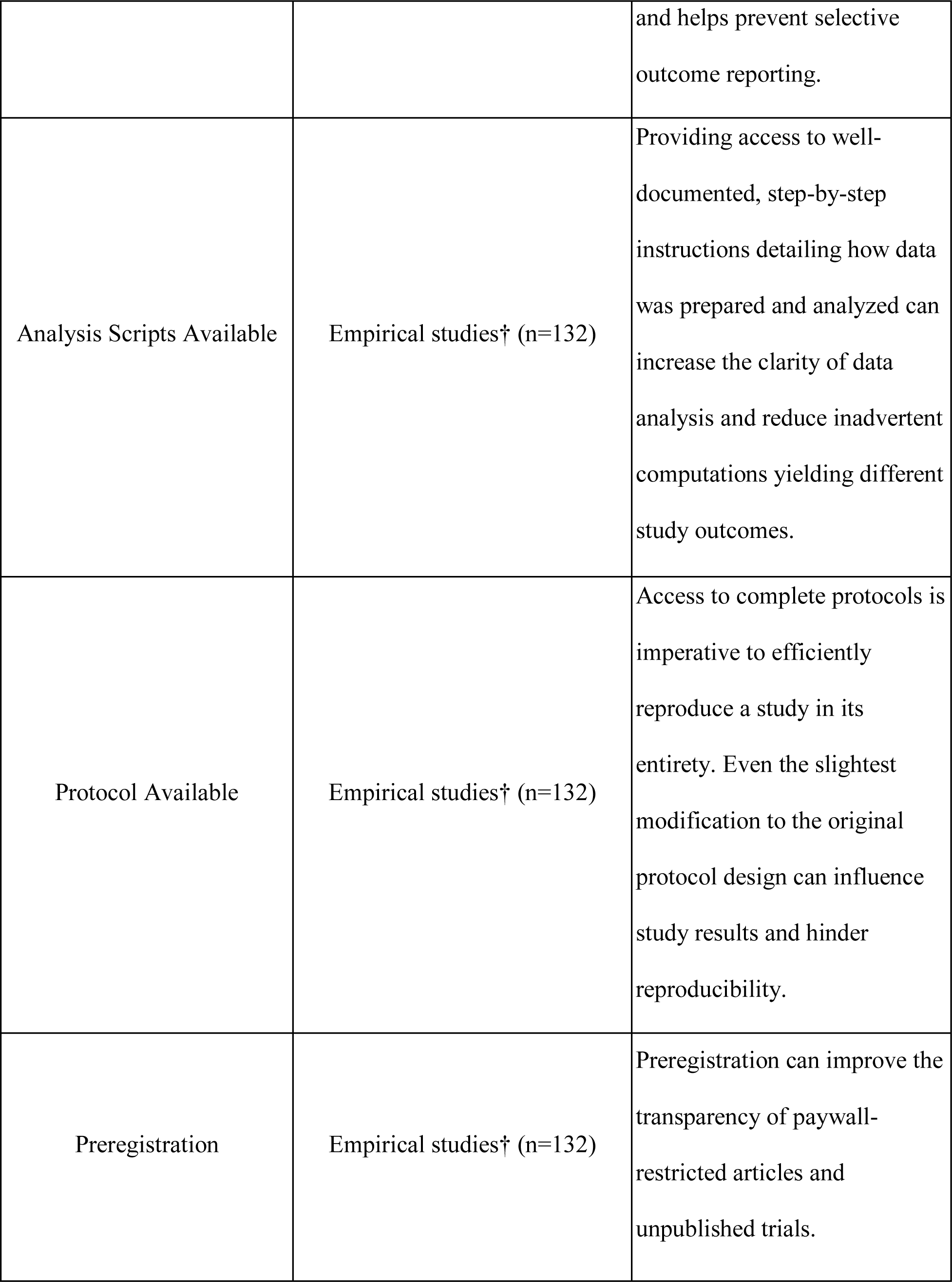
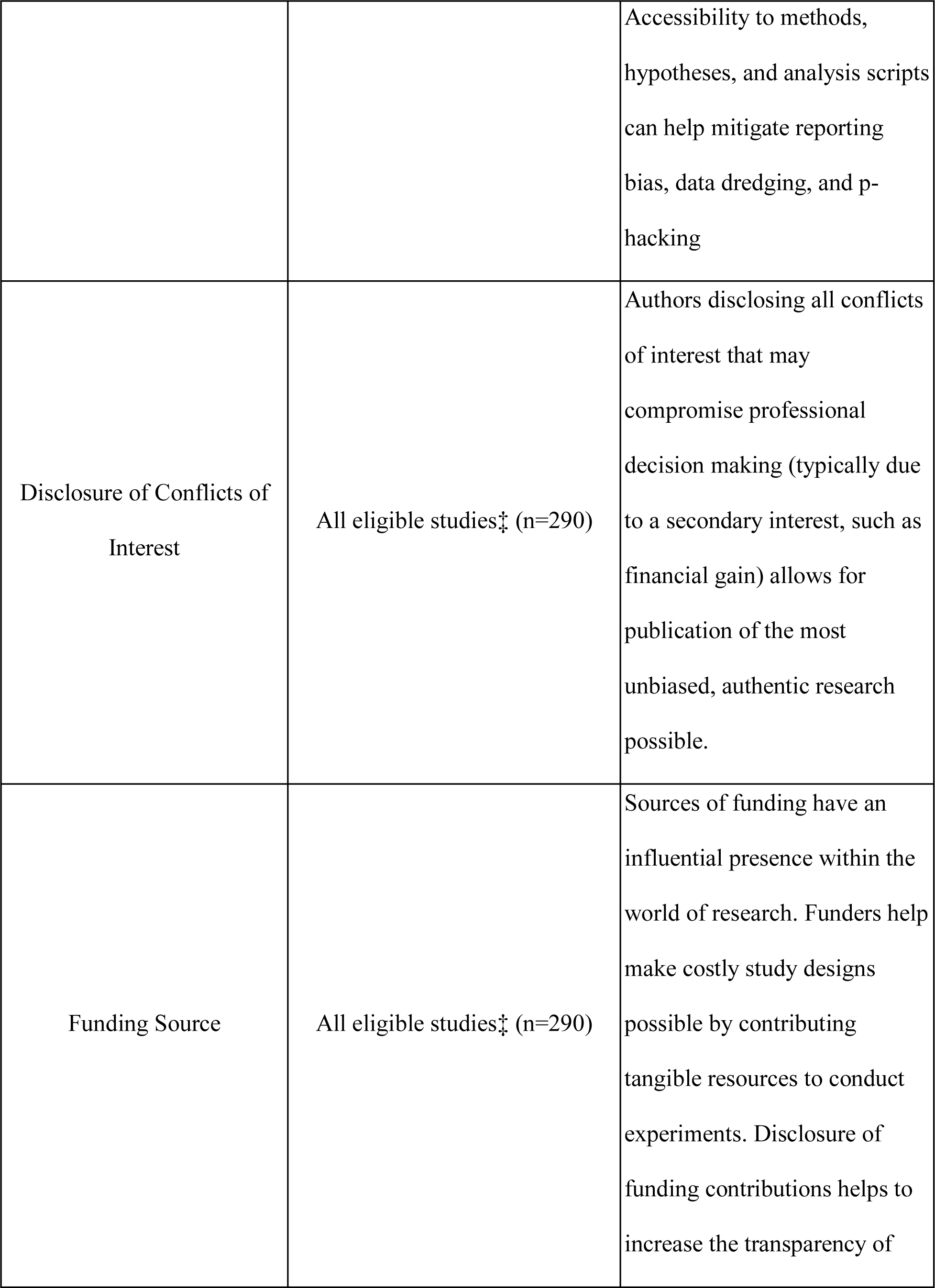
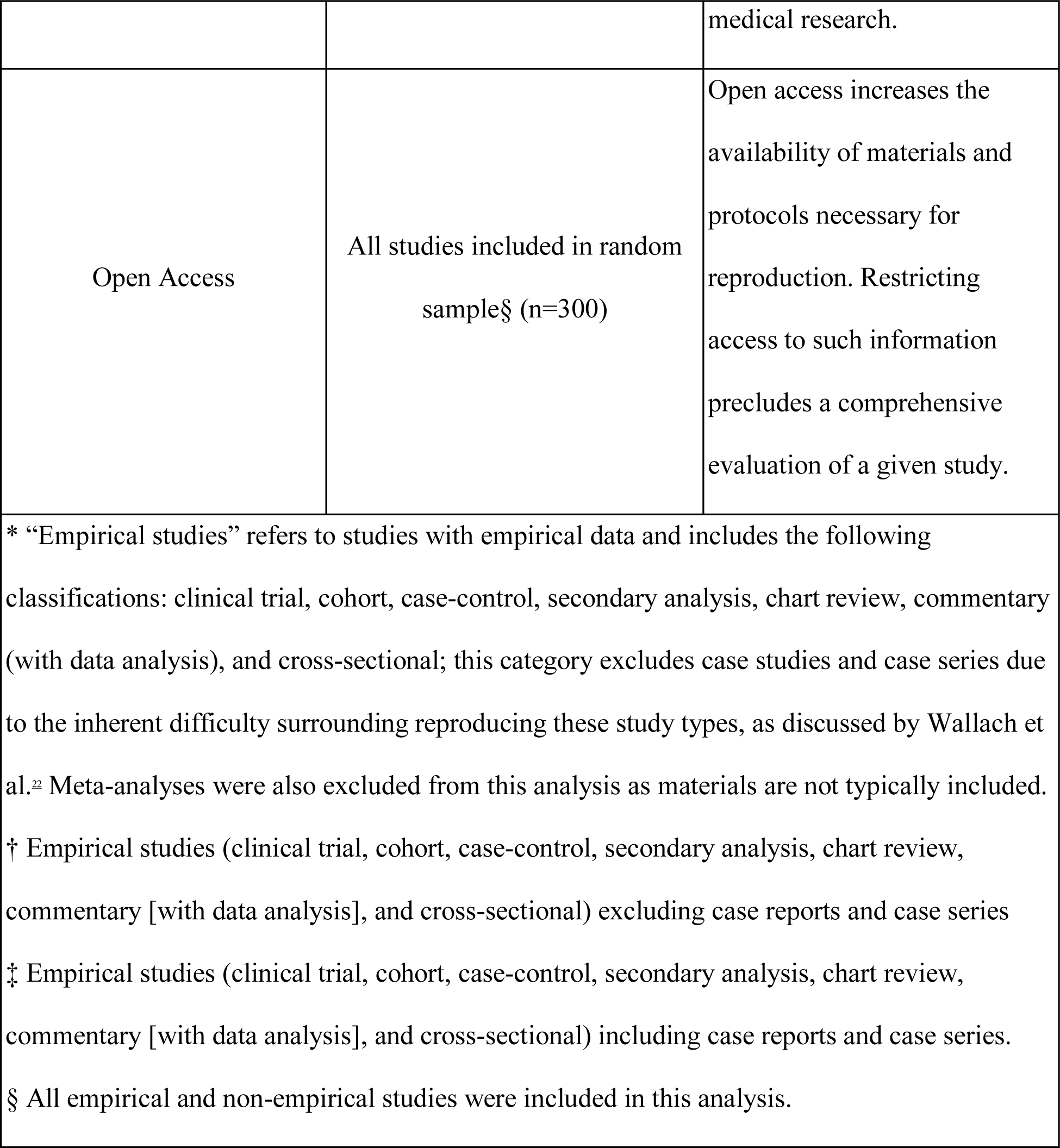
Indicators of Reproducibility. Analysis of variables within each publication was dependent on study classification. Rationale detailing study classification is available here: https://osf.io/x24n3/

| **Indicators of Reproducibility and Transparency** | **Study Types Included for Analysis of Reproducibility Indicator** | **Usefulness for Reproducing Medical Literature** |
| --- | --- | --- |
| Materials Available | Empirical studies* (n=127) | Providing all materials, including stimuli, survey instruments, and computer code/software used for data collection or running experiments increases the feasibility by which researchers' are able to successfully replicate a study using identical methodology. |
| Raw Data | Empirical studies† (n=132) | Disseminating records of collected information in its unaltered, digital form facilitates validation of study outcomes through reanalysis |
|  |  | and helps prevent selective outcome reporting. |
| Analysis Scripts Available | Empirical studies† (n=132) | Providing access to well-documented, step-by-step instructions detailing how data was prepared and analyzed can increase the clarity of data analysis and reduce inadvertent computations yielding different study outcomes. |
| Protocol Available | Empirical studies† (n=132) | Access to complete protocols is imperative to efficiently reproduce a study in its entirety. Even the slightest modification to the original protocol design can influence study results and hinder reproducibility. |
| Preregistration | Empirical studies† (n=132) | Preregistration can improve the transparency of paywall-restricted articles and unpublished trials. |
|  |  | Accessibility to methods, hypotheses, and analysis scripts can help mitigate reporting bias, data dredging, and p-hacking |
| Disclosure of Conflicts of Interest | All eligible studies‡ (n=290) | Authors disclosing all conflicts of interest that may compromise professional decision making (typically due to a secondary interest, such as financial gain) allows for publication of the most unbiased, authentic research possible. |
| Funding Source | All eligible studies‡ (n=290) | Sources of funding have an influential presence within the world of research. Funders help make costly study designs possible by contributing tangible resources to conduct experiments. Disclosure of funding contributions helps to increase the transparency of |
|  |  | medical research. |
| Open Access | All studies included in random sample§ (n=300) | Open access increases the availability of materials and protocols necessary for reproduction. Restricting access to such information precludes a comprehensive evaluation of a given study. |
| * “Empirical studies” refers to studies with empirical data and includes the following classifications: clinical trial, cohort, case-control, secondary analysis, chart review, commentary (with data analysis), and cross-sectional; this category excludes case studies and case series due to the inherent difficulty surrounding reproducing these study types, as discussed by Wallach et al.<sup>22</sup> Meta-analyses were also excluded from this analysis as materials are not typically included.<br>† Empirical studies (clinical trial, cohort, case-control, secondary analysis, chart review, commentary [with data analysis], and cross-sectional) excluding case reports and case series<br>‡ Empirical studies (clinical trial, cohort, case-control, secondary analysis, chart review, commentary [with data analysis], and cross-sectional) including case reports and case series.<br>§ All empirical and non-empirical studies were included in this analysis. |  |  |

**Supplemental Table 2.**
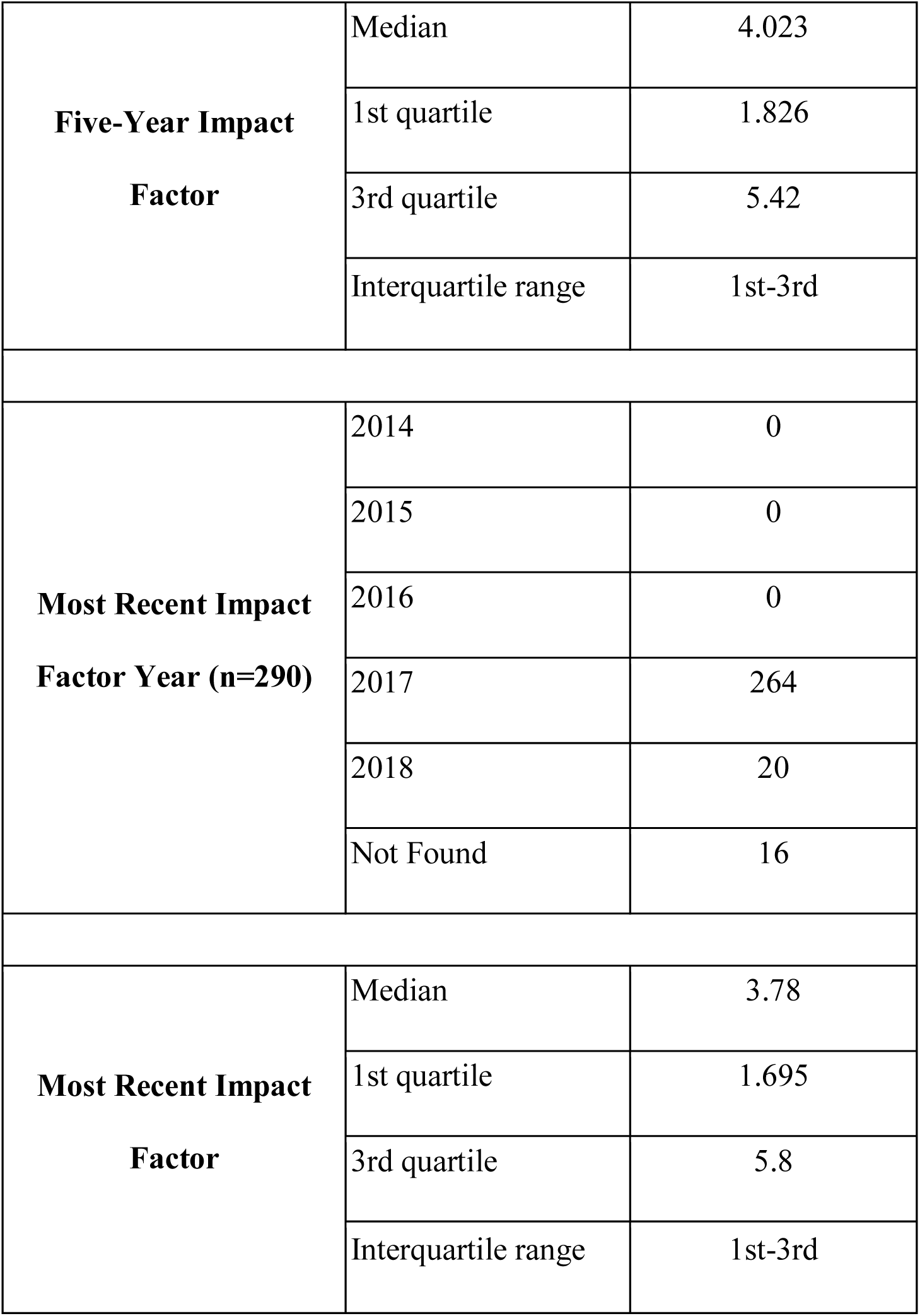
Cardiovascular System Journal Impact Factor.

|  |  |  |
| --- | --- | --- |
| **Five-Year Impact**<br>**Factor** | Median | 4.023 |
|  | 1st quartile | 1.826 |
|  | 3rd quartile | 5.42 |
|  | Interquartile range | 1st-3rd |
| **Most Recent Impact**<br>**Factor Year (n=290)** | 2014 | 0 |
|  | 2015 | 0 |
|  | 2016 | 0 |
|  | 2017 | 264 |
|  | 2018 | 20 |
|  | Not Found | 16 |
| **Most Recent Impact**<br>**Factor** | Median | 3.78 |
|  | 1st quartile | 1.695 |
|  | 3rd quartile | 5.8 |
|  | Interquartile range | 1st-3rd |

**Supplemental Table 3.**
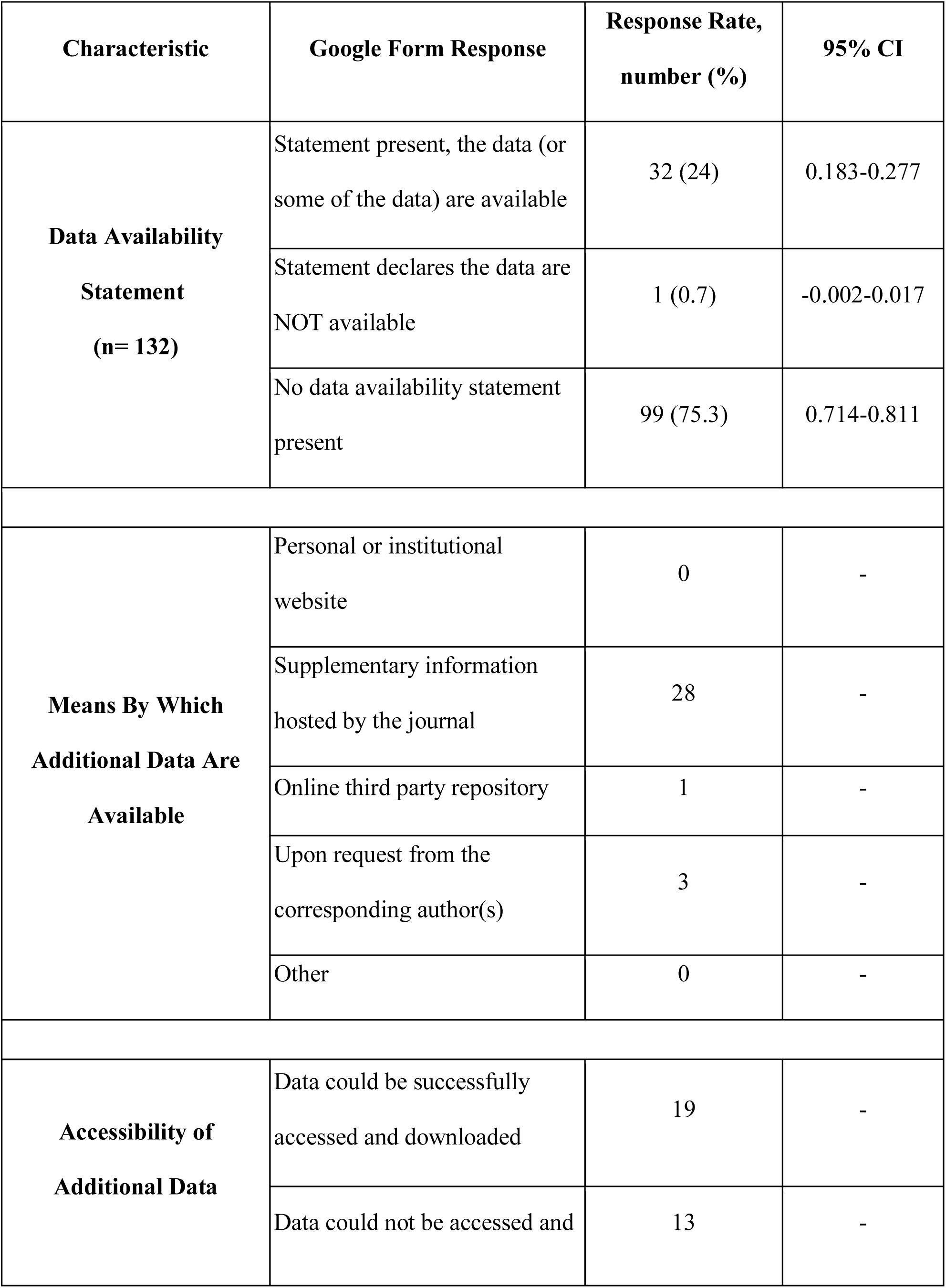
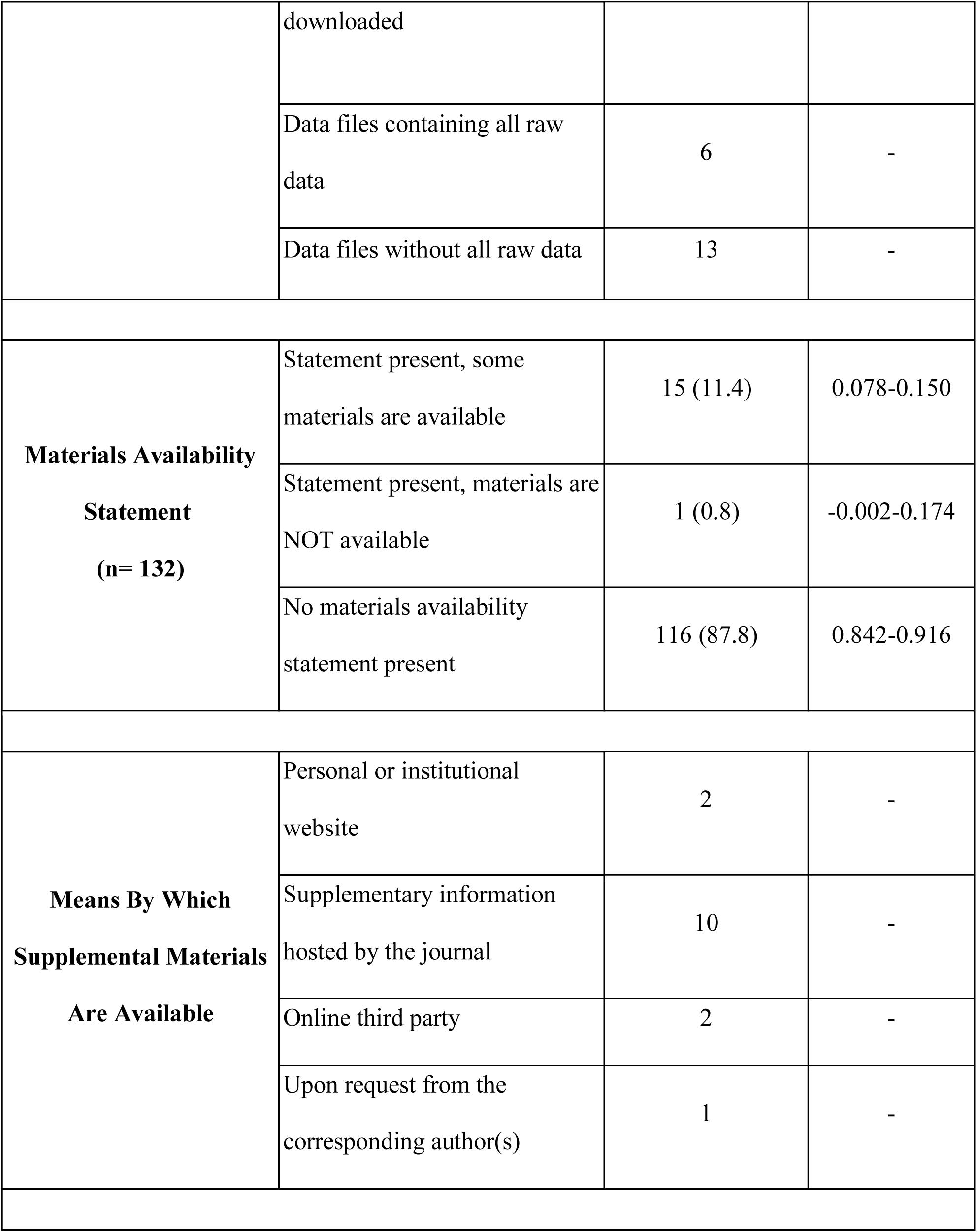
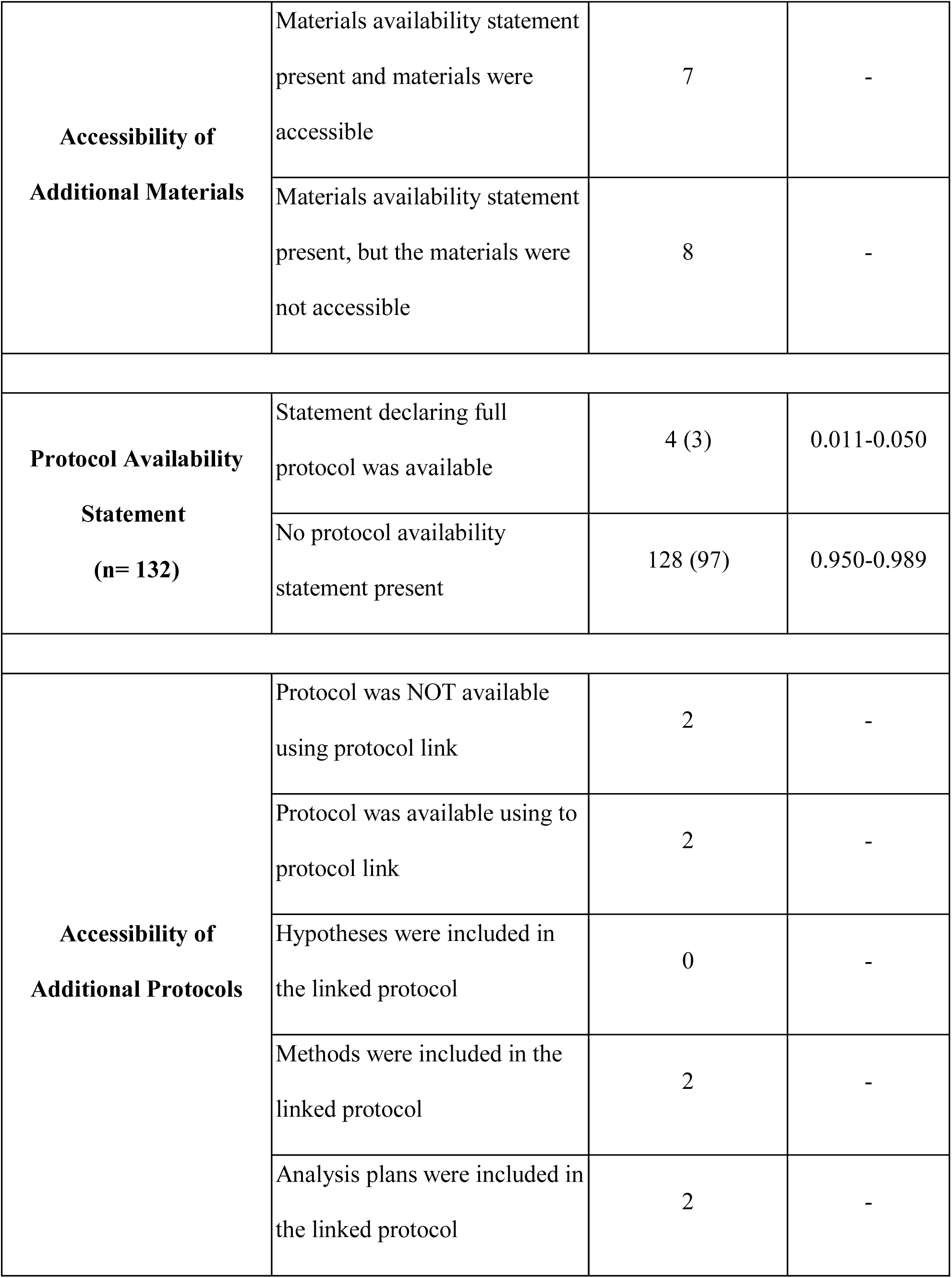
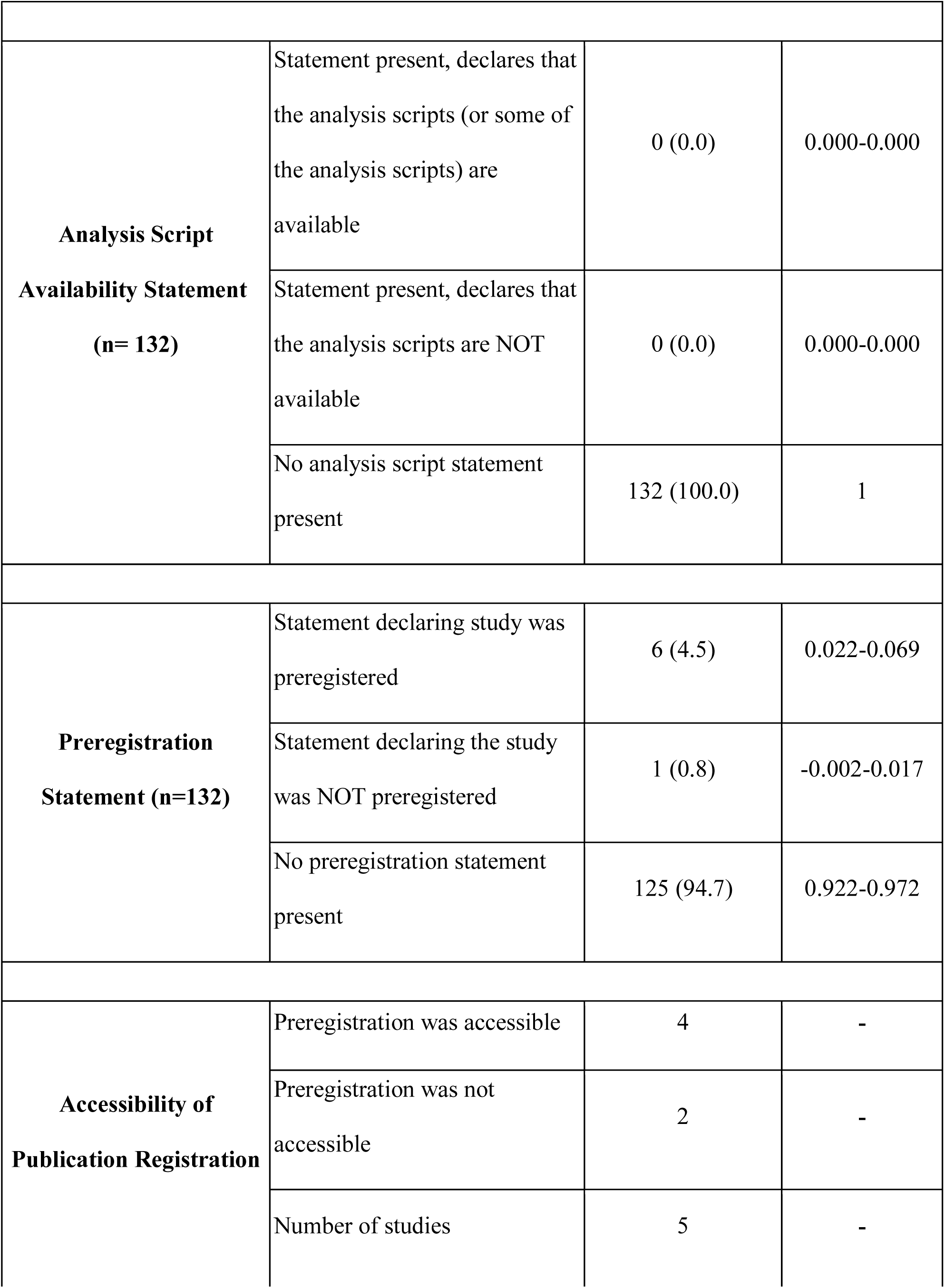
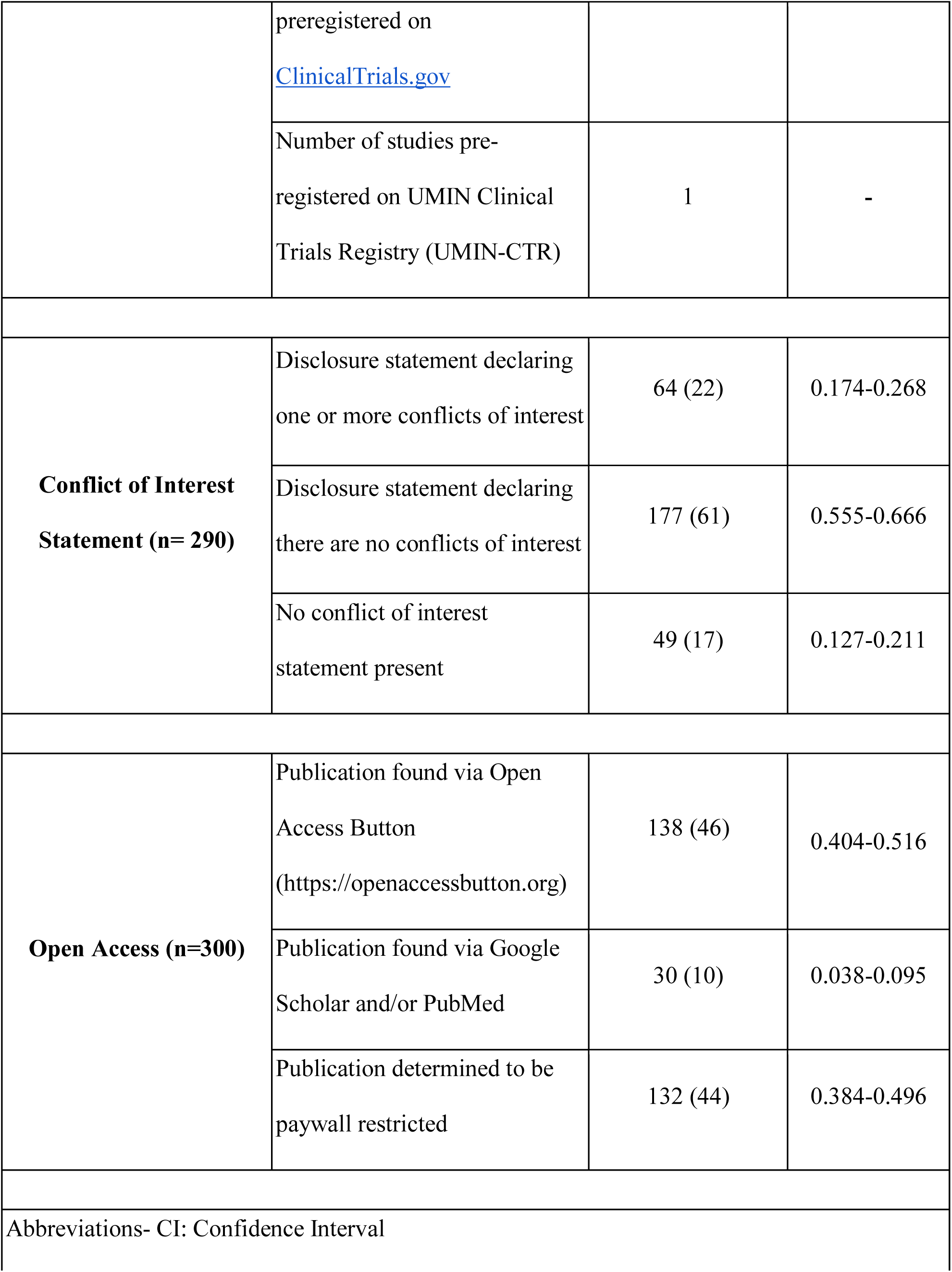
Additional Reproducibility and Transparency Characteristics of Publications in Cardiology Journals.

**Supplemental Table 4.** Number of Times Publications Has Been Cited in a Meta-Analysis and/or Systematic Review Article.

|  | **n (%)** |
| --- | --- |
| **No citation** | 106 (80.3) |
| **A Single Citation** | 20 (15.2) |
| **One to Five Citations** | 5 (3.8) |
| **Greater Than Five Citations** | 1 (0.7) |

